# Environmental monitoring and health assessment in an industrial town in central India: A cross-sectional study protocol

**DOI:** 10.1101/2022.02.07.22270576

**Authors:** Tanwi Trushna, Vikas Dhiman, Satish Bhagwatrao Aher, Dharma Raj, Rajesh Ahirwar, Swasti Shubham, Subroto Shambhu Nandi, Rajnarayan R Tiwari

**Author notes:** Corresponding Author1: Dr Subroto Shambhu Nandi [ ] Postal Address with Contact details: Dr. S. S. Nandi, Department of Environmental Exposure Assessment and Monitoring (Air), ICMR-National Institute for Research in Environmental Health, Bhopal Bypass Road, Bhauri-462030, Madhya Pradesh, India. Corresponding Author 2: Dr. Tanwi Trushna [ ] Postal Address with Contact details: Dr. Tanwi Trushna, Department of Environmental Health and Epidemiology, ICMR-National Institute for Research in Environmental Health, Bhopal Bypass Road, Bhauri- 462030, Madhya Pradesh, India.

## Abstract

**Background:** Textile industry has been widely implicated in environmental pollution. The health effects of residing near manufacturing industries are not well documented in India, especially in central India. Hence, a cross-sectional environmental monitoring and health assessment study was initiated as per directions of the local authorities.

**Methods:** Comprehensive exposure data about the concentrations of relevant pollutants in the ambient air and ground water samples in the study area will be collected over 1 year. Using stratified random sampling, 3003 apparently healthy adults will be selected from the study area. Sociodemographic and anthropometric information, relevant medical and family history, and investigations including spirometry, electrocardiogram, neurobehavioral tests, and laboratory investigations (complete blood count, lipid profile and random blood glucose) will be conducted. Finally Iodine azide test and heavy metal level detection in urine and blood samples respectively will be conducted in a subset of selected participants to assess individual pollution exposure. Ethics approval has been obtained from the Institutional Ethics Committee of the National Institute for Research in Environmental Health (No: NIREH/IEC-7-II/1027, dated 07/01/2021).

**Discussion:** This manuscript describes the protocol for a multi-disciplinary study that aims to conduct environmental monitoring and health assessment in residential areas near viscose rayon and associated chemical manufacturing industries. Although India is the second largest manufacturer of rayon, next only to China, and viscose rayon manufacturing has been documented to be a source of multiple toxic pollutants, there is a lack of comprehensive information about the health effects of residing near such manufacturing units in India. Therefore implementing this study protocol will aid in filling in this knowledge gap.

## Introduction

Industrial development has contributed to economic growth but at the same time industrial activities constitute a major source of environmental pollution.^1^ Adverse health effects posed by industrial pollution is well-established.^1^ De Sario et al.,2018 in their scoping review identified 762 epidemiological studies which conclude that living in industrial areas is associated with a wide range of diseases ranging from malignancies and respiratory diseases to adverse birth outcomes and even premature mortality.^2^ Of these studies, few were conducted in India that focussed on the public health and environmental effects of industrial emission. Chatham-Stephens et al., 2013 using the Toxic Sites Identification Program developed by the Blacksmith Institute and the United Nations Industrial Development Organization identified 221 point-sources of pollution from industrial activities in India that pose risk to public health.^3^ The authors calculated that exposure to pollutants generated in industrial regions in developing countries like India, Indonesia, and the Philippines pose equal or even greater risk to human health than widespread communicable diseases like malaria and account for almost 8 million years of healthy life lost due to disease, disability or premature death (i.e. 828,722 Disability-adjusted Life Years) among the 8.6 million exposed population in the three countries.^3^ Therefore, research is needed to assess the environmental health effects of major industries in India.

India, being a rapidly developing economy, is home to multiple industries. Textile industry, plays a critical role in Indian economy as it is the second largest source of employment for a country which is counted amongst the top producers, exporters as well as consumers of textiles in the world.^2^ However, the textile industry has been widely implicated in pollution of various environmental matrices like air, water and soil both in India^3,4^ and other countries.^5,6^ It accounts for almost twenty percent of global industrial activity induced water pollution^5^ and is associated with emission of many gaseous pollutants into ambient air.^7^ Multiple studies have documented the adverse health effects of exposure to the chemical pollutants that emanate from textile and associated chemical manufacturing.^8,9^ In India, research on the health effects of such textile industry has mostly been focussed on individuals who are occupationally exposed to the chemical pollutants.^10–12^ The health effects of residing near manufacturing industries are not as well documented.

The industrial area of Birlagram Nagda in the central Indian state of Madhya Pradesh, has been under scrutiny due to the pollution emanating from the multiple viscose rayon textile and associated chemical manufacturing industries operating in the area.^13^ As per data compiled by the Central Pollution Control Board (CPCB), the apex government run agency monitoring environmental pollutant concentrations in the country, the area has a comprehensive environment pollution index (CEPI) score of 66.67, which indicates that it is among the “severely polluted” industrial clusters in India.^14^ Major chemical pollution associated with viscose rayon textile manufacturing occur secondary to gaseous emission of carbon disulphide (CS_2_), Hydrogen Disulphide (H_2_S), Hydrogen Chloride (HCl), Chlorine (Cl_2_), Sulphur oxides (SO_2_ and SO_3_) and untreated discharge of viscose fibre wastewater effluent containing high concentration of heavy metals, and other toxic substances into natural water sources.^7^ However, there is hardly any scientific evidence regarding the health effects of residing near the industries of Birlagram. Considering the growing health concerns of the local public, regulatory authorities in India directed the industries operational in the region to implement pollution control strategies as well as commissioned detailed environmental and health-oriented investigation of the issue.^15^ The current study was initiated as per directions of the local authorities to evaluate the health effects of exposure to multiple pollutants emitted from the industrial activity in Birlagram. Specifically, the objectives of the current study include:

i. To measure the concentration of gaseous pollutants (i.e. CS_2_, H_2_S, HCl, Cl_2_, and SO_2_) in the ambient air in the study area.
ii. To measure the concentrations of heavy metals and anions [i.e. lead (Pb), mercury(Hg), aluminium (Al), chloride (Cl^-^) and sulphate(SO_4_ ^2-^)] in ground water samples.
iii. To determine the association between the concentration levels of pollutants in air and water samples with that in bio-samples (blood and urine).
iv. To identify the association between exposure to pollutants and health effects in study participants.

## Methods and analysis

### Study Design

This is the protocol for a cross-sectional research study that aims to collect and correlate comprehensive exposure and health outcome information to ascertain objective achievement (see Fig 1). This study will be conducted during a duration of 1 year. The time schedule of various study methods of this protocol is described in Fig 2.

**Fig 1:**
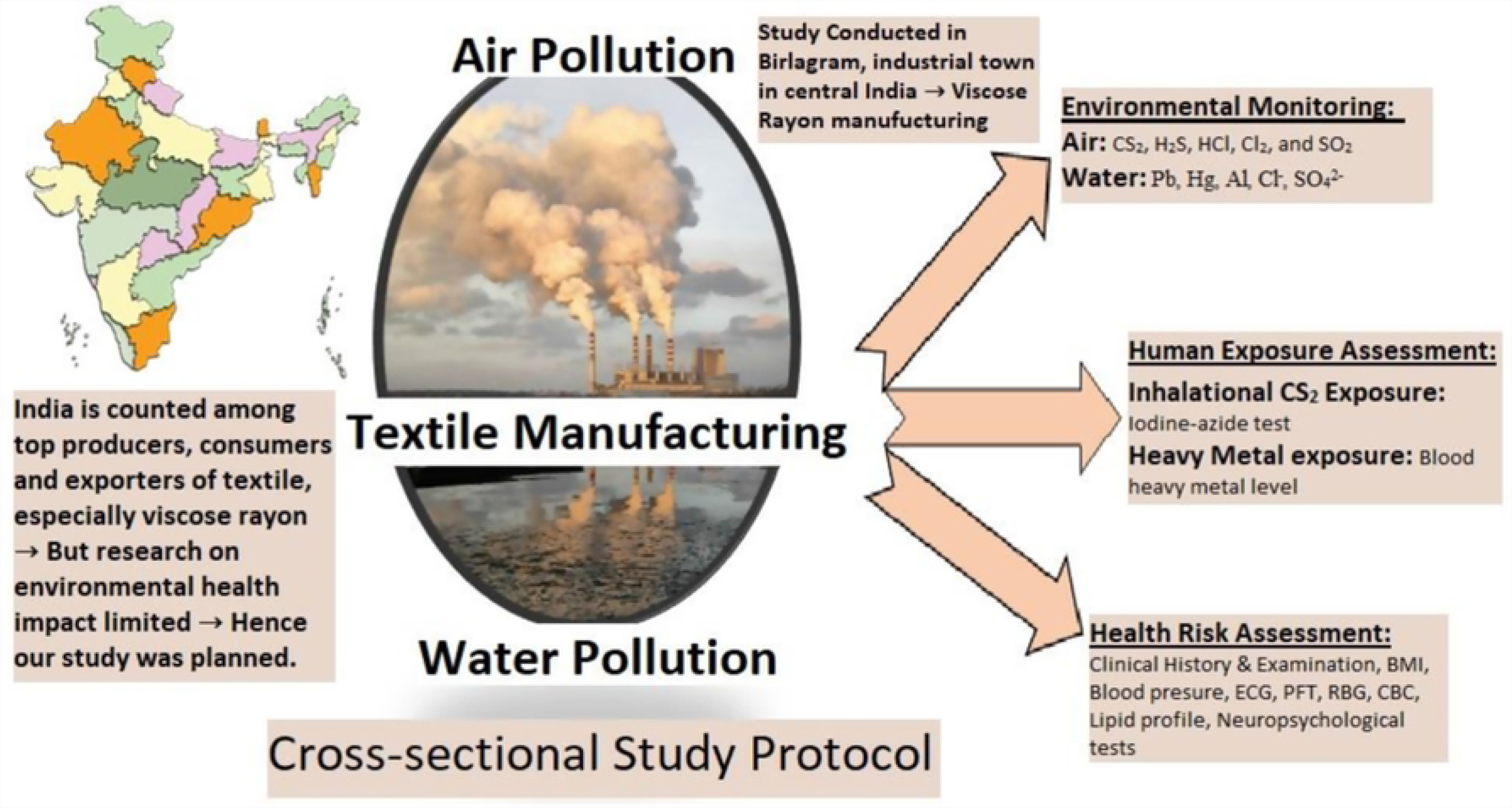
Graphical abstract: Overview of the study methodology [Note: carbon disulphide (CS_2_), Hydrogen sulphide (H_2_S), Hydrogen Chloride (HCl), Chlorine (Cl_2_), Sulphur oxides (SO_2_), lead (Pb), mercury(Hg), aluminium (Al), chloride (Cl^-^), sulphate (SO_4_ ^2-^), body mass index (BMI), Electrocardiogram (ECG), Pulmonary function test (PFT), Random Blood glucose (RBG), Complete blood count (CBC)] (The graphical abstract image has been created by the research team using copyright-free images of Indian map and industry from https://pixabay.com)

**Fig 2:**
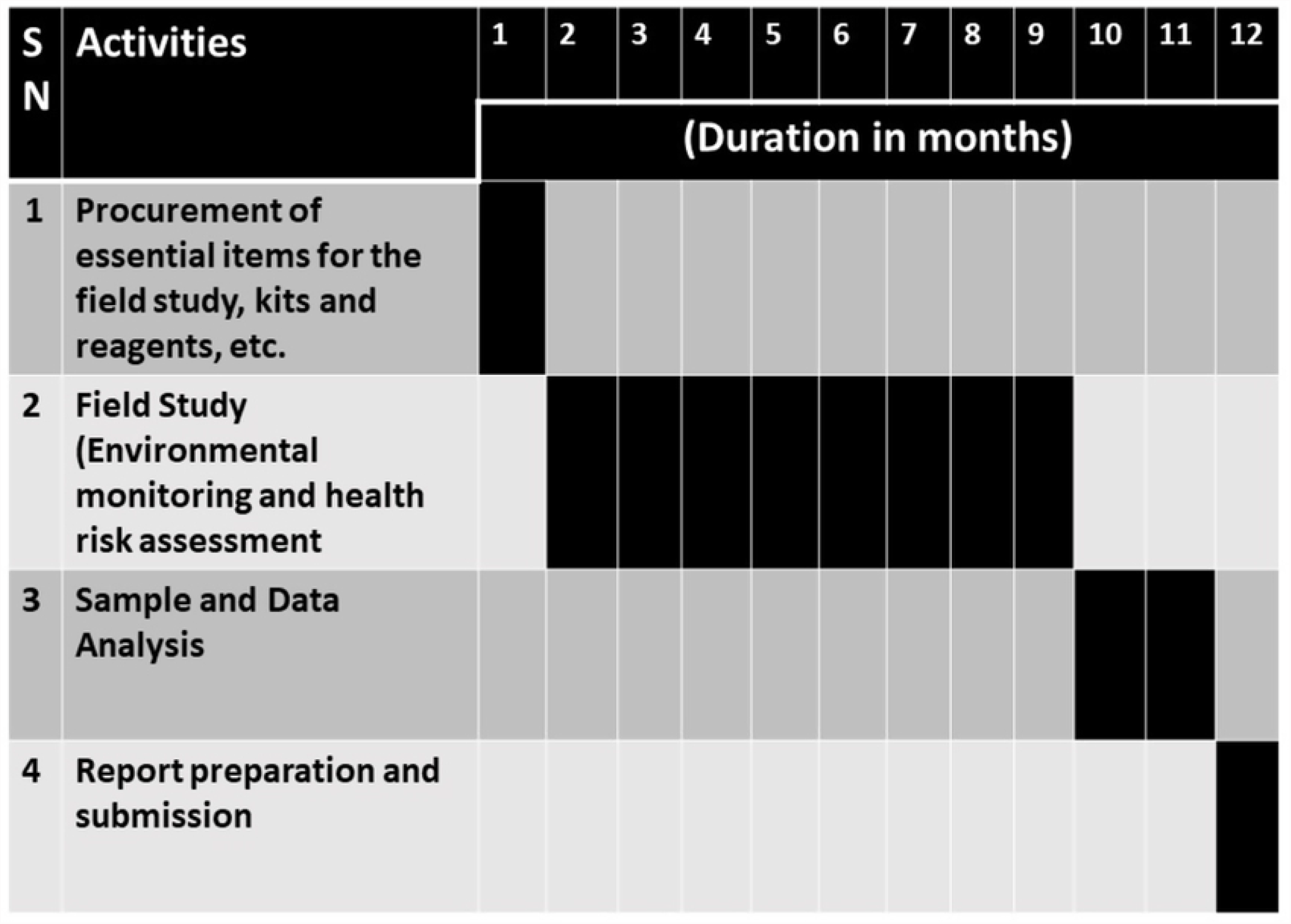
Time Schedule of Activities Note: Each numbered column represents one month of the project duration with the total duration of the project being 12 months.

### Setting

This study will be conducted in and around the Birlagram industrial region located in Nagda city in the Ujjain district (administrative unit) in the central Indian state of Madhya Pradesh (MP). Almost 72 million individuals reside in MP,^16^ most (75%) of whom reside in rural areas and depend on agriculture for their livelihood.^17^ However, industrial activity in MP is gradually increasing^18^ and multiple large scale industries in the Ujjain district^19^ constitute an emerging public health concern for the province that needs to be evaluated.

The Birlagram industrial area is situated within the borders of Nagda city which in turn is surrounded on all sides by villages.^13^ The industries situated in Birlagram are directly or indirectly (through production of chemical raw materials) involved in synthetic textile (viscose rayon) manufacturing.^20^ Including the population residing in Birlagram itself, Nagda city has a total population of more than a million. River Chambal is the main source of water for the region and previous research findings highlight the poor quality of its water.^21^ Details on air quality of the area are lacking in published literature.

### Participant selection and sampling strategy

#### Sample size calculation

As the health effects of residing near textile manufacturing industries in Birlagram, India are not previously documented, we calculated the sample size using the statistics reported by a previous study conducted in Taiwan where authors found that 26.3% of the population exposed to emissions from a viscose rayon manufacturing industry had electrocardiogram (ECG) abnormalities.^22^ Since one of our outcomes is identification of the proportion of exposed population in Birlagram, India with ECG abnormalities, we calculated sample size as follows:

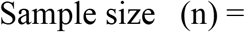

Where, p= Expected population proportion= 0.26; q= 1-p = 0.74; z= z value for 5% level of significance is 1.96; R= Non-response rate (assumed to be 30% as pooled non-response rate reported by a systematic review of participation response in Indian industrial surveys^23^); Deff= Design effect(1.25 for systematic sampling); and d= Margin of error 2% (assumed to be 0.02). The calculated sample size was 3003.

#### Sampling strategy

In this study stratified random sampling technique will be used for selection of participants where stratification will be done based on environmental factors that are likely to affect exposure. Since both distance from fixed contamination source as well as local environment features such as meteorology, topography, etc. are the major factors affecting the dispersion of both ambient air as well as water pollutants,^24^ a map of the study area was created to assist in selection of exposure monitoring locations and in participant sampling (see Fig 3). Briefly, taking Birlagram industrial region as the centre, villages located within consecutive circles of radius 2km, 5km and 10km were demarcated on the map using ArcInfo (version 10). This strategy was used as it is expected that gaseous pollutants’ dispersion occurs homogenously in all directions independent of anthropogenic roadways and that with increase in distance the pollutant concentration will become diluted enough to prevent adverse health effects. ^25^

**Fig 3:**
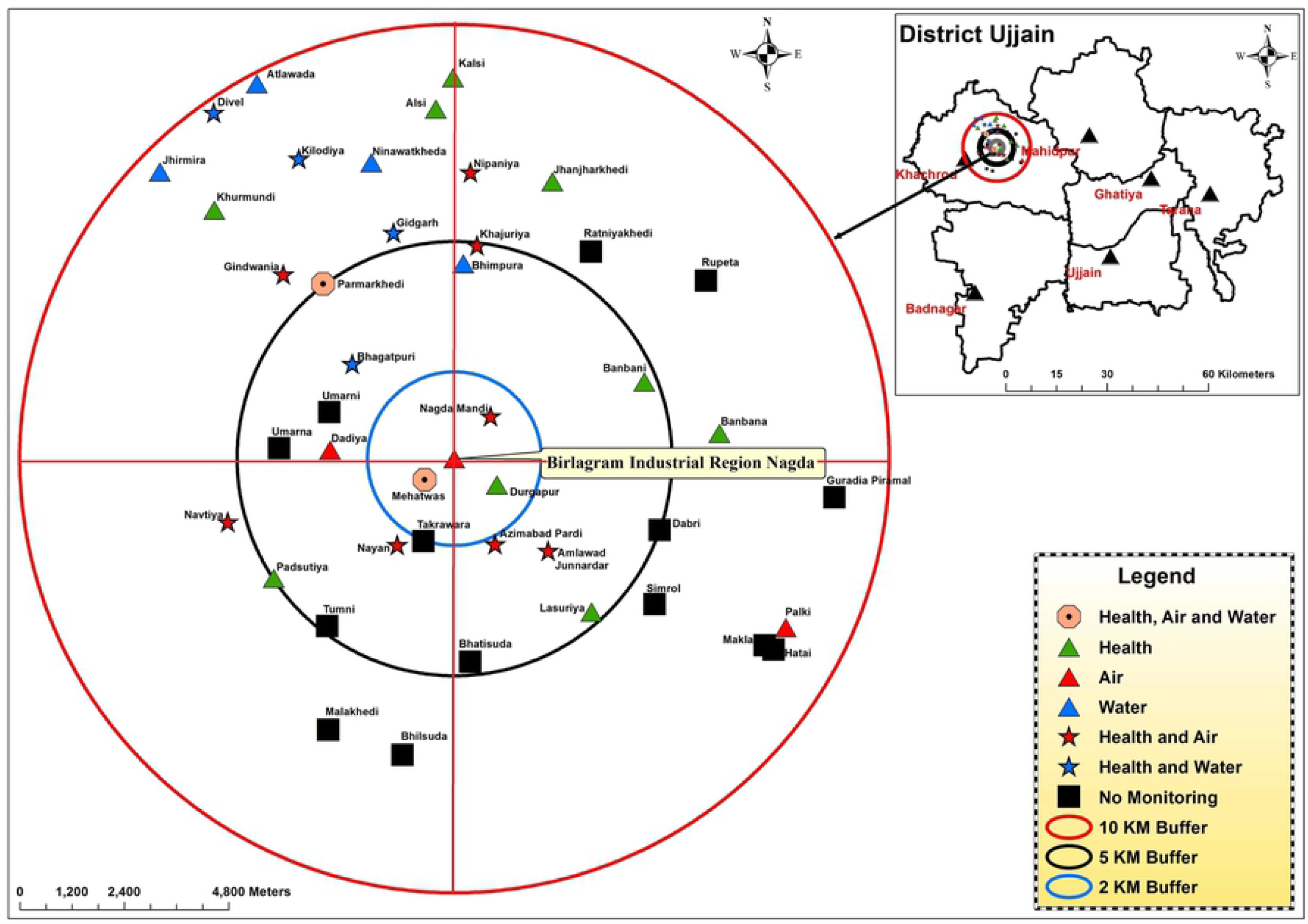
Map of the study area created by research team to assist in selection of exposure monitoring locations and in participant sampling. [Note: The inset in the map shows the Ujjain district of the central Indian province of MP. Within the Khachrod Tehsil of the Ujjain district, lies the study area of the current project. Considering the Birlagram industrial region as the centre, three concentric circular regions were demarcated based on distance (2, 5 and 10 kilometers from the centre).]

Based on seasonal wind pattern data retrieved from local authorities, the north-eastern quadrant of the circular study area was seen to be the downwind area for most of the year. The downstream direction of River Chambal falls in the north-western quadrant of the circular study area. Therefore the majority of the pollutants are expected to disperse in the northern direction increasing the potential risk faced by the residents and so, we decided to over-represent the northern hemisphere.

Finally, 2000 participants will be selected from the villages in the northern hemisphere while 1003 will be taken from the villages in the southern hemisphere within the 10km radius comprising a total of 3003 participants. In each hemisphere, a fixed number of villages will be selected randomly from those situated in each of the 0-2km, 2-5km and 5-10km zones. Further in each selected village, using the list of all individuals residing in the villages that will be retrieved from the local authorities as the sampling frame, individuals meeting the eligibility criteria will be randomly invited for participation in the study. Total number of individuals sampled in each village will be proportionate to the total population of the selected village.

Participants will be selected for the study only if they fulfill all the following criteria:

- Adults (≥18 years of age) of both genders (except pregnant women),
- Should have been residing for at least 1 year in the selected village,
- No history of neuropsychiatric conditions that might hinder completion of neurobehavioral tests,
- No history of other conditions that are contraindications for performing PFT like presence of any active disease (e.g. pulmonary Tuberculosis, active haemoptysis, orofacial pain, acute respiratory infection including COVID-19) or recent history (within previous 1 month) of myocardial infarction/aneurysm/surgery, etc.

### Environmental monitoring of pollutants at study sites

Pollutants for assessment in both environmental and human matrices were selected based upon their relevance to viscose rayon and associated chemical manufacturing industry identified through review of previously published literature.^7^

#### Monitoring of gaseous pollutants in ambient air

The ambient air monitoring will be carried out in the study area (see figure 2) for identified gaseous pollutants viz., CS_2_, H_2_S, HCl, Cl_2_, and SO_2_using a factory calibrated, electrochemical sensor based multi-gas monitor (Make: Swan**®** Environmental Pvt. Ltd. India, Model: GRI-IAT; Sensor make: Membrapor**®**, Switzerland). The monitoring will be carried out in post-monsoon season (October-January). The instrument has an inbuilt geographical positioning system (GPS) to record the coordinates of sampling site. The ambient air monitoring will be carried out continuously for 24 hours in at identified sites. In case of cluster of sites, a representative site will be identified for gaseous monitoring. The monitor will be placed at more than 3 metres height from ground level at a place having uninterrupted AC power supply. The data loggers present in the instrument will store the data at set frequency and will be utilized for further statistical analysis. The sampling and monitoring of ambient air for said pollutants will be carried out following the existing guidelines of Central Pollution Control Board (CPCB), Ministry of Environment, Forests and Climate Change (MOEFCC), New Delhi, India.^28^

#### Monitoring of metallic pollutants in groundwater

Representative groundwater samples (120 mL) from sources including tube-wells, hand-pumps and open wells will be collected from the selected downstream villages (Figure 2) located along the bank of the Chambal river to identify the presence and concentration of heavy metals such as lead, mercury and aluminium and anions such as chloride and sulphate. Groundwater samples will be collected from all available and accessible sources in each village. Samples will be collected in pre-cleaned high density polypropylene (HDPE) bottles from various sources after removing stagnant water using appropriate purging methods.^27^ Also, a few water samples will be collected from the Chambal River at specified locations to assess metallic pollutants. Collected water samples will be labeled and transported to the laboratory at room temperature for initial measurement of physiological parameters including pH, total dissolved salts (TDS), conductivity, and salinity. Subsequently, the samples will be preserved with trace metal grade nitric acid (pH <2) and stored at 4 °C until analysis.

Analysis of toxic metals, viz. Pb, Hg and Al in the collected groundwater samples will be carried out using inductively coupled plasma optical emission spectroscopy (iCAP**®** 7400 Duo ICP-OES, ThermoFisher Scientific**®** Pvt Ltd) using the EPA method 200.7 (revision 4.4)^28^. Calibration standard for each element will be prepared from multi-element stock solutions (1000 mgL^-1^) in triple distilled water. Detection of Pb and Al will be performed using standard sample introduction setup, whereas for Hg, the hydride generation sample introduction system will be used. Data acquisition will be done using the Qtegra**®** ISDS Software at interference free wavelengths for these elements. Detection of chloride and sulphate will be carried out using titration methods.

### Human Exposure Assessment

Following biomonitoring will be done to assess exposure in the selected participants:

#### Inhalational CS_2_ Exposure

Iodine-azide test will be carried out for estimation of carbon disulfide exposure of the participants. The iodine-azide test is based on the fact that certain constituents in the urine of persons exposed to carbon disulphide catalyze the reaction between iodine and sodium azide.^29,30^

Participants will be asked to provide approx. 50 mL of clean catch spot urine sample in a sterile container which will be stored at -20 °C till analysis. Preparation of reagents and buffers as well as the test procedures will be carried out as previously described.^29,31^ The exposure coefficient will be calculated to estimate Carbon disulfide exposure.

#### Metal (Pb, Hg and Al) exposure

To assess blood levels of afore-mentioned heavy metals, 1 mL of whole blood will be digested with 4 m concentrated hydrochloric acid (HCl) and 2 mL hydrogen peroxide (H_2_O_2_) in the microwave digestion system (digestion cycle: heating to 200 °C, maintain for 15 minutes, cooling to 50 °C). The digested samples will be diluted to 35 mL for analysis of Pb, Al and Hg using the ICP-OES method as outlined in the above section.

### Health Assessment

Following health-related data will be collected from all selected participants:

#### Questionnaire-based collection of data on demography, exposure and lifestyle

A structured questionnaire which has been developed by the researchers will be pilot-tested and thereafter wards used for collection of socioeconomic and demographic details along with information regarding lifestyle factors, occupational and relevant exposure history. The questionnaire will be administered by trained investigators in the local language.

#### Anthropometry and Blood pressure monitoring

Height and weight will be measured using stadiometer and weighing scale, respectively and will be used to calculate body mass index (BMI). For Indian population, BMI 18.5–22.9 kg/m^2^ will be considered as normal, 23–24.9 kg/m^2^ as overweight and more than or equal to 25 kg/m^2^ will be considered as obesity.^32^ Blood pressure will be measured as per the International Society of Hypertension’s 2020 Global Hypertension Practice Guidelines^33^ in sitting position on the left arm using validated electronic blood pressure monitor (Omron**®**), after 5 minutes of rest. Systolic and diastolic blood pressure will be recorded as the mean of last two of three readings taken 1 minute apart. Hypertension will be defined as the study participants having raised systolic and/or diastolic blood pressure (more than or equal to 140 and/or 90 mm Hg, respectively). ^32^

#### Clinical History and Examination

The information regarding current and previous cardiological, respiratory and neurological symptoms will be taken by the physicians from the participants using a clinical proforma. The details regarding the onset, duration, severity of symptoms, diurnal variation, aggravating/relieving factors, and associated symptoms details will be noted. A general physical examination and detailed respiratory, cardiological and neurological examination of all participants will be conducted which will include higher mental function examination, speech assessment, cranial nerve examinations, examination of the motor system and muscle power, examination of the sensory system, reflexes and coordination, and gait assessment, etc. All abnormal findings will be recorded in a pre-designed the clinical proforma.

#### Neurobehavioral Tests

CS_2_, mainly used in the production of viscose rayon fibre, is one of the main air pollutant in Nagda Industrial town. Although overt CS_2_-induced neurotoxicity is no longer a serious problem nowadays, neurobehavioral problems due to chronic exposure to low levels of CS_2_ are not uncommon.^36^ Previous studies, although mostly conducted in occupational set-up, have shown problems in cognition,^34,35^ fine coordination,^36^ learning and memory,^37^ visual attention & concentration,^38^ sensory and motor functions^39^ and other behavioral functions^40^ in humans due to CS_2_ exposure. Hence a battery of neurobehavioral tests (see Table 1) will be employed in the present study to screen any known neuro-behavioral effects due to CS_2_ exposure.

**Table 1:**
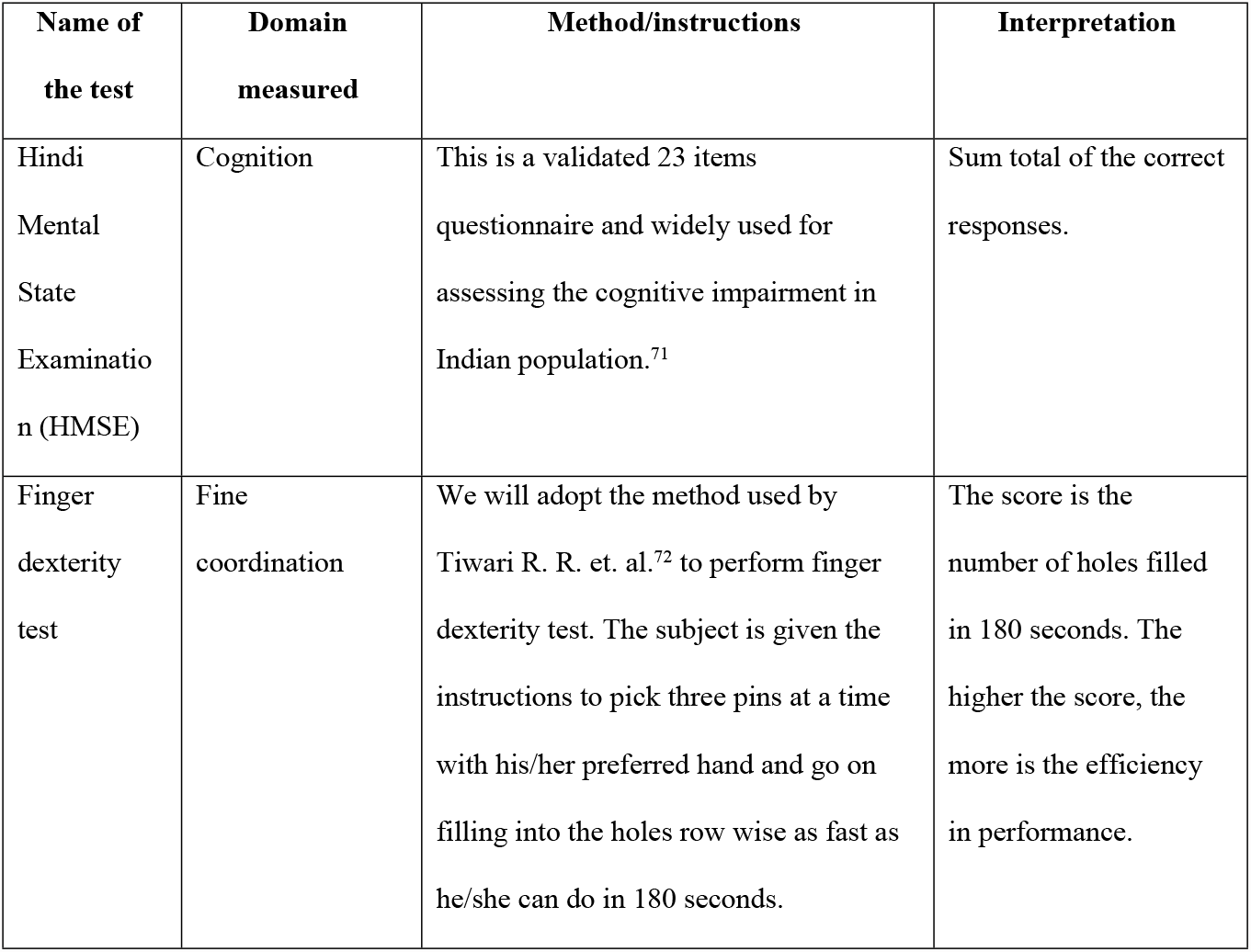
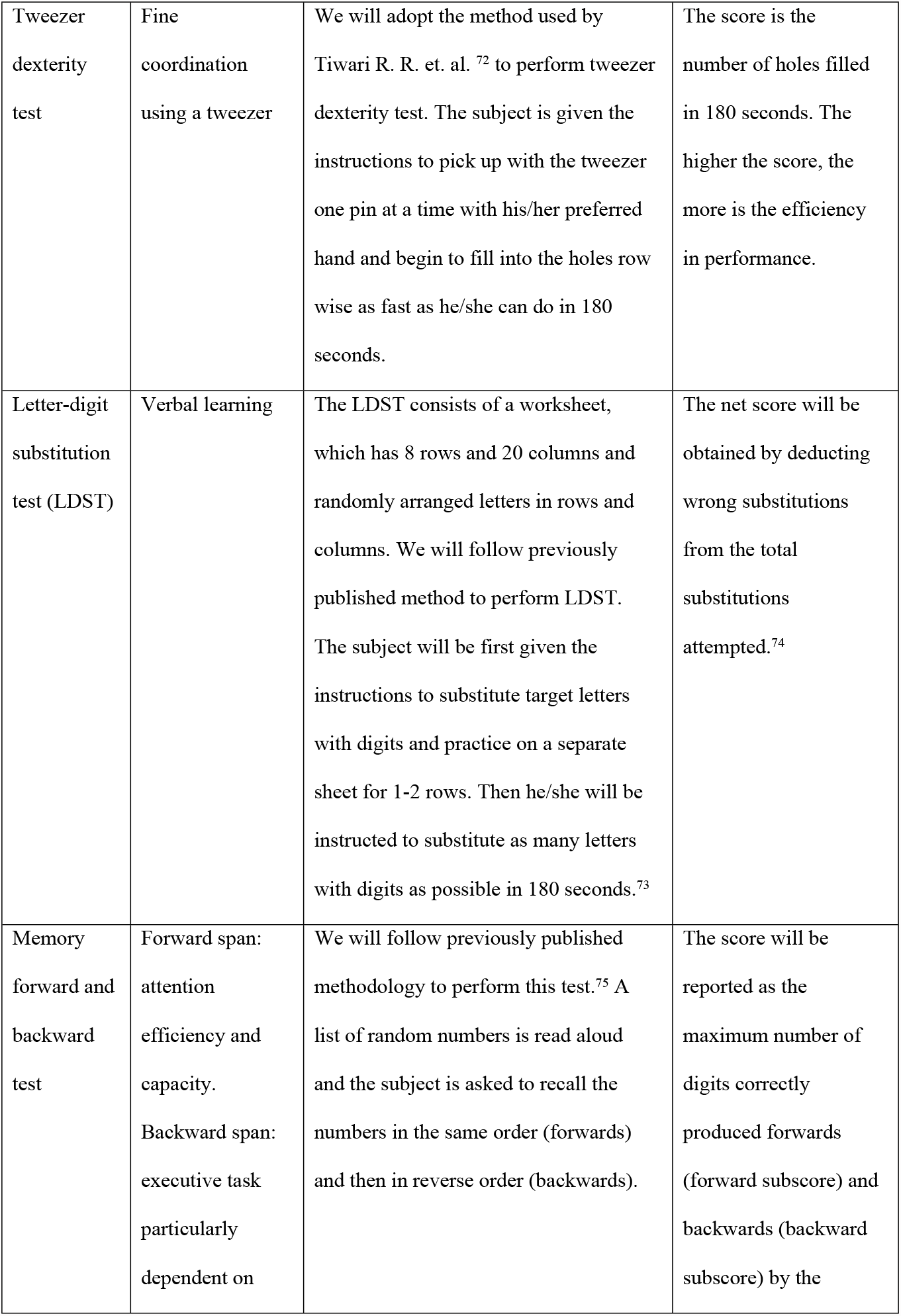
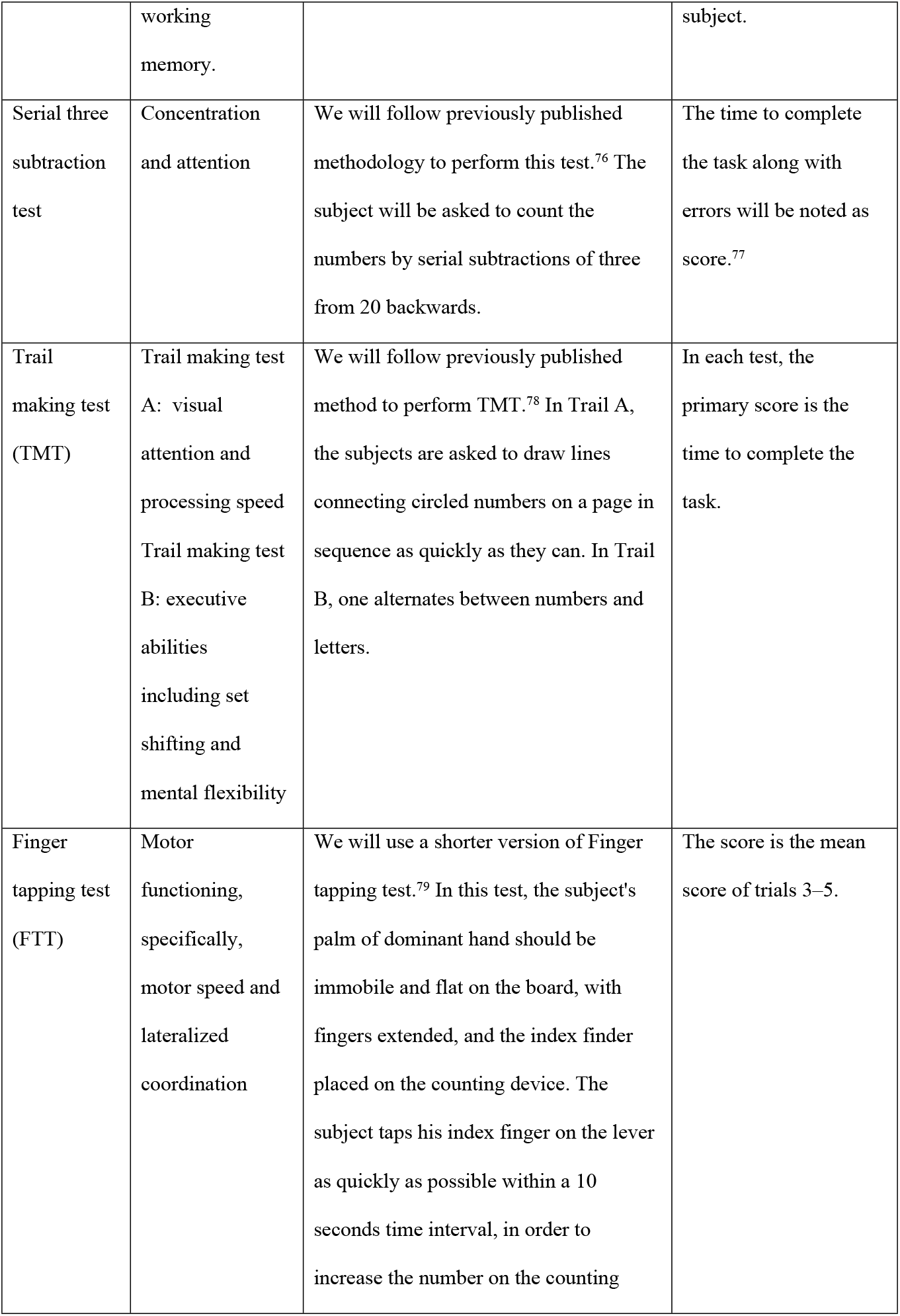
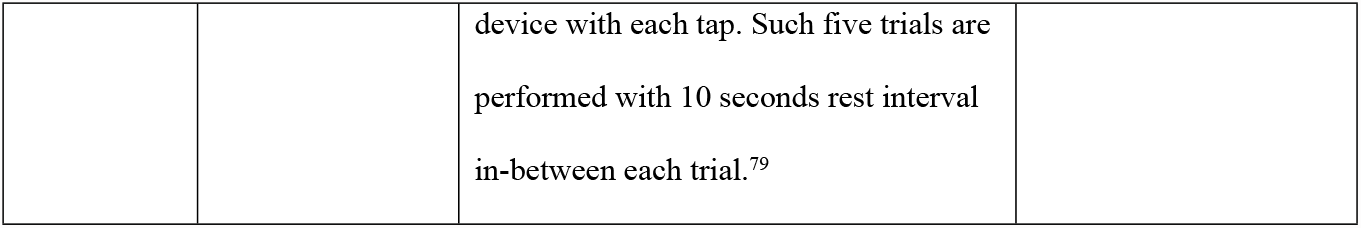
Details of neurobehavioral tests to be administered to study participants.

#### Clinical Investigations

##### i. Spirometry

Pulmonary Function testing will be conducted using portable spirometer (Cosmed**®** Pony FX) on comfortably seated participants by a trained technician, supervised by a trained physician, following national guidelines,^50^ to measure forced expiratory volume in 1st second (FEV1), forced vital capacity(FVC), and peak expiratory flow (PEF). Of three acceptable spirograms defined as the difference between the two largest FVC and the two largest FEV1 measurements are less than or equal to 0.150 litres, the best value for each parameter will be included for analysis.

##### ii. Electrocardiography (ECG)

A portable 12-lead Electrocardiogram device (BPL CardiArt**®** 9108D) will be used to measure electrical activity of the heart following American Heart Association guidelines.^51^ Briefly, in a private screened area, ECG recording will be done by previously trained medical staff on the chest and limbs of supine and relaxed participants. Trained physicians will interpret the ECG recording as per standard guidelines.^52^

##### iii. Laboratory investigations

A battery of clinical laboratory investigations comprising of Random Blood Glucose, Complete Blood Counts, and Lipid profile (Total cholesterol, HDL cholesterol, LDL cholesterol, Triglyceride) will be carried out for all participants of the study. Blood sample collection will be carried out by trained technician according to standard operating procedure in compliance with WHO Best Practices.^53^ To undertake the afore-mentioned three tests, briefly, a total of 5mL sample of venous blood will be collected from the median cubital vein of the participants by trained technician maintaining sterile precautions. 2 mL blood will be collected in EDTA vacutainer for Complete blood count which will be processed within 4 hours using a fully automated hematology analyzer (Sysmex**®**). 3 mL blood sample will be collected in plain vacutainer from which serum will be separated and stored at -20 °C till analysis. The serum will be used for estimation of lipid profile (Total cholesterol, HDL, LDL, Triglyceride). Lipid profile will be estimated using semi-automated biochemistry analyzer (ERBA**®**). Random blood glucose will be estimated on the spot from a drop of blood at the time of sample collection, using a hand held glucometer (Accu-Chek**®**).

### Quality control

Necessary measures as per the National guidelines for data quality in surveys^48^ such as data collection by trained researchers, validation and pilot-testing of study instruments prior to use, meticulous checking and standardization of equipment, and reagents to be used, etc. will be taken to maintain data quality at the highest possible level.

### Data management and analysis

All participants’ data will be coded by assigning unique identifiers. These codes will be used to identify and link various parameters of the individual participants. Data from the filled questionnaires/clinical proforma as well as the laboratory analysis values will be entered into the latest version of Microsoft Excel spreadsheets. Statistician of the team will supervise data entry and to ensure error-free data at least 10 % of the entered data will be randomly verified.

Once data entry is complete, hard copies of questionnaire/proforma will be stored in a secure archive in the institute. Similarly, all entered data will be stored in password protected computer systems with access restricted to the research team alone. The primary investigator will be in-charge of data safety and back-up maintenance.

All statistical analyses will be performed using IBM SPSS Statistics (version 25). For ordinal and continuous variables, the values will be given as descriptive statistics (like mean, median, standard deviation, 95% confidence interval, interquartile range); and categorical variables will be given as numbers (percentages) and proportions. Association for different health aspects with pollution level of various parameters of air/water will be made using Pearson’s chi-square test for independent categorical data. By considering the variability within and between groups; t-test and ANOVA will be used to analyze the mean differences between two and more groups respectively for various anthropological parameters, the concentration of air/water pollutants, etc. The Mann –Whitney U test and Wilcoxon signed ranks test will be used for independent and dependent for ordinal and continuous data w.r.t. different health aspects. Pearson and Spearman’s correlation coefficient will be used to examine the association between different variables. Multiple linear and logistic regression models will be used to estimate the relationship for different neurological assessments with a set of the independent variable. For all statistical tests, two-sided P-values <0.05 will be considered to be significant.

### Ethics and Dissemination

Ethics approval has been obtained from the Institutional (Human) Ethics Committee, National Institute for Research in Environmental Health (No: NIREH/IEC-7-II/1027, dated January 7 2021). Before the initiation of research activities, local authorities of the selected village will be approached for consent and aid in logistics. Prior to the start of data collection written informed consent will be taken from each participant.

Dissemination of the study findings will be done through submission of study technical report to the funding agency, distribution of health reports to all the participants while maintaining their confidentiality and finally, through publications.

## Discussion

This manuscript describes the protocol for a cross-sectional study being conducted to comprehensively assess the health effects of residing near viscose rayon and associated chemical manufacturing industries. Viscose rayon is one of the oldest man-made commercial fiber^55^ that is still quite popular today with almost 4.5 million tons being manufactured annually worldwide^56^. It is manufactured by chemically dissolving natural cellulose, commonly in the form of wood pulp,^55^ using a Sodium hydroxide (NaOH) /carbon disulfide (CS_2_) dissolution system.^56^ However, during this process almost 25-30% of the CS_2_ used is not recovered and escapes as emission into the ambient air.^56^ In their review, Jiang et al., 2020 report that for every ton of viscose produced, almost 20kg of exhaust and 300-600 tons of waste water containing heavy metals and other toxic residues are generated.^56^ Our study will produce up-to-date evidence specific to the Indian context which is especially relevant considering the fact that India is the second largest manufacturer of rayon, next only to China.^55^

Chronic low dose exposure to CS_2_, a major component of the viscose rayon industry exhaust, affects human physiology especially the cardiovascular and nervous system.^8,57,58^Studies have documented abnormalities in blood pressure,^59^ ECG findings,^60^and other blood parameters like total cholesterol, HDL, LDL and triglycerides.^61^Similarly CS_2_ exposure has been linked to peripheral neuropathy,^62^abnormalities in colour vision^63^ and neurobehavioral pattern.^64^Though fewer studies have been done to assess the effects of CS_2_ on the respiratory system, Spyker et al. in 1982 showed alterations in pulmonary function.^65^ Effect on eyes, reproductive system and renal system have also been noted, although following prolonged heavy exposure.^39^ Other gaseous pollutants emitted during viscose rayon and associated chemical manufacturing like H_2_S, HCl, Cl_2_, and SO_2_ are known to affect human health, particularly the cardiorespiratory and nervous system.^66–68^

Similarly, water and soil contamination occurring due to effluents being discharged by viscose rayon manufacturing industries has been reported in literature.^69^ Effluents of textile industries using high amounts of salts may promote groundwater salinization due to the high mobility of salts/ions in the soil, and thus deteriorate drinking and irrigation water quality that may impact human health and land fertility/ crop productivity.^70,71^ The viscose fibre wastewater has also been shown to have a wide variety of residues ranging from heavy metals to cellulose and lignin remnants, most of which resist biodegradation.^7,72^ Previously published studies report exposure of workers in viscose rayon industries to heavy metals like mercury, lead, etc.^73^ Chronic heavy metal exposure is again known to cause multiple adverse cardiorespiratory and neurological effects.^74^

Most of the evidence regarding health effects of exposure to pollution emanating from viscose rayon and associated chemical manufacturing originates from outside India with there being hardly any published epidemiological evidence focussing on either workers in Indian industries^75^ or nearby residents. However, the difficulties perceived by the residents living in Nagda industrial region has been scientifically documented^13^ as well as has been highlighted in multiple other communication media^76^. Previous reports by local regulatory authorities showed substantial heavy metal contamination of groundwater in and around Nagda region.^15^ Furthermore, the chemical industries in Nagda region are engaged in production of various chlorinated and sulphur-containing product (e.g., poly-aluminium chloride, bleaching powder, calcium chloride, chloromethane, chlorosulphonic acid)^77^ which can potentially lead to introduction of other heavy metals like Aluminium and ions like Cl^-^and SO_4_ ^-^ ions into the surrounding groundwater. Therefore, this study is planned to collect pertinent and timely comprehensive information on the combined effect of the multiple toxic gases including H_2_S, Sulphur oxides, HCl, Cl_2_, etc. emanating as exhaust from the viscose rayon and associated chemical manufacturing industries as well as assess the physiological parameters and heavy metal/ ion concentration in the groundwater in the study area.

Industrial pollution is a multidimensional issue since it occurs in multiple environmental matrices and in turn affects multiple organ systems in the human. Therefore, in our study we will focus on multiple objectives with an overarching goal to assess the potential health effects of exposure. A major strength of this study is therefore its multidisciplinary research team with expertise of investigators ranging from health sciences like epidemiology and neurology to environmental sciences and basic science. To strengthen the association between pollutant concentration in environmental matrices and health effects, human exposure assessment will be done in a subset of the population to ascertain dose-response relation. Therefore, exposure to CS_2_ will be confirmed using the urine azide test which has been reported to adequately reflect occupational CS_2_ inhalation by published literature.^31–33^ Furthermore, blood levels of heavy metals will also be determined to estimate exposure of villagers to toxic metals, viz. Pb, Al and Hg from various environmental matrices, viz. air, water and food/soil. However, pertaining to resource limitation, biomonitoring will be conducted in a subset of the total study population.

The current study also faces certain limitations. Long term follow-up of the participants to establish a temporal relationship between exposure and outcome is not possible in our cross-sectional study. Considering the ongoing dispute between local residents, viscose rayon manufacturers and the local authorities, the current study prioritised timely delivery of baseline health effect information. Future research might therefore focus on longitudinal data collection on the environmental health effects of viscose rayon manufacturing. Furthermore, past exposure assessment will be done through questionnaire data and is therefore, prone to recall bias. Although data will be collected for multiple relevant confounders including other environmental/meteorological, socioeconomic and diet/lifestyle factors, the risk of residual confounding cannot be completely eliminated.

## Data Availability

No datasets were generated or analysed during the current study. All relevant data from this study will be made available upon study completion.

## Abbreviations

Al: Aluminium
ANOVA: Analysis Of Variance (Statistical Test)
BMI: Body Mass Index
CEPI: Comprehensive Environment Pollution Index
Cl^-^: Chloride Ion
Cl_2_: Chlorine Gas
COVID-19: Coronavirus Disease Caused By The SARS-Cov-2 Virus
CPCB: Central Pollution Control Board
CS_2_: Carbon Disulphide
EDTA: Ethylene diamine tetra acetic Acid
ECG: Electrocardiogram
FEV1: Forced Expiratory Volume In 1st Second
FVC: Forced Vital Capacity
H_2_S: Hydrogen Disulphide
Hcl: Hydrogen Chloride
Hg: Mercury
HDL: High-Density Lipoprotein
LDL: Low-Density Lipoprotein
MP: Madhya Pradesh
Pb: Lead
PFT: Pulmonary Function Test
pH: Potential Of Hydrogen (Measure Of Acidity/Basicity)
PEF: Peak Expiratory Flow
SO_2_: Sulphur Dioxide
SO_3_: Sulphur Trioxide
SO_4_^2-^: Sulphate Ion
TDS: Total Dissolved Salts
°C: Degrees Celsius
Km: Kilometer
Kg/m^2^: Kilogram Per Square Metre
mL: Milliliter
mgL^-1^: Milligram Per Litre
mm Hg: Millimetre Of Mercury

## Acknowledgements

We would like to acknowledge the technical and field staff of ICMR-NIREH, participants as well as the local authorities and health workers working in the villages where we conducted our exposure assessment and health outcome assessment surveys. We would also like to thank Dr. Vivek Parashar, Research Officer, R.D. Gardi Medical College Ujjain, Madhya Pradesh, India, for his support in creating the study area maps.

## Authors’ contributions

All authors met the authorship criteria set forth by the International Committee for Medical Journal Editors and read and approved the final manuscript

Conceptualization: SN, RRT, RA, VD, and TT. Data curation: DR. Formal analysis: Not applicable (This is study methodology manuscript). Funding acquisition: SN, RRT. Methodology: RRT, SN, SS, RA, VD, TT, SA, and DR. Project administration: SN, RRT. Visualization: TT, DR. Writing–original draft: TT. Writing–review & editing: SN, SS, RA, VD, SA, DR, and RRT.

## Availability of data and materials

Not applicable (Data sharing does not apply to this article which describes a study protocol and thus no datasets have been analyzed yet).

## Ethics approval and consent to participate

Ethics approval has been obtained from the Institutional (Human) Ethics Committee, National Institute for Research in Environmental Health (No: NIREH/IEC-7-II/1027, dated January 7 2021). Written informed consent with permission for publication will be obtained from all participants.

## Consent for publication

Not applicable for this manuscript since no identifying images or other personal/clinical details of participants that can compromise anonymity have been presented in this protocol manuscript.

## References

1. Rahman MM, Alam K, Velayutham E. Is industrial pollution detrimental to public health? Evidence from the world’s most industrialised countries. BMC Public Health. 2021 Jun 18;21(1):1175

2. Dixit P, Lal RC. Inclusive Growth & Social Responsibility - A Critical Analysis of IndianTextile Industry. MERC Global’s International Journal of Management. 2019;7(2):202–10

3. Bhatia D, Sharma NR, Kanwar R, Singh J. Physicochemical assessment of industrial textile effluents of Punjab (India). Applied Water Science. 2018 May 22;8(3):83

4. Manikandan P, Palanisamy PN, Baskar R. Physicochemical analayis of textile industrial effluents from Tirupur city, TN, India. undefined [Internet]. 2015 [cited 2021 Sep 27]; Available from: https://www.semanticscholar.org/paper/PHYSICO-CHEMICAL-ANALYSIS-OF-TEXTILE-INDUSTRIAL-TN%2C-Manikandan-Palanisamy/f7b02c20e197ce3bde89d48a197365d62ab869e7

5. Aldalbahi A, El-Naggar ME, El-Newehy MH, Rahaman M, Hatshan MR, Khattab TA. Effects of Technical Textiles and Synthetic Nanofibers on Environmental Pollution. Polymers. 2021;13(1)

6. Shirvanimoghaddam K, Motamed B, Ramakrishna S, Naebe M. Death by waste: Fashion and textile circular economy case. Science of The Total Environment. 2020 May 20;718:137317

7. Madhav S, Ahamad A, Singh P, Mishra PK. A review of textile industry: Wet processing, environmental impacts, and effluent treatment methods. Environmental Quality Management. 2018 Mar 1;27(3):31–41

8. Pappuswamy M, Sundaram R, Kuppanna HM, Periyasamy T. Carbon Disulfide (CS2) Induced Chromosomal Alterations and Apoptosis in Circulated Blood Lymphocytes of Personnel Working in Viscose Industry. Asian Pacific Journal of Cancer Biology. 2018 Feb 25;3(1):17–24

9. Sieja K, von Mach-Szczypiński J, von Mach-Szczypiński J. Health effect of chronic exposure to carbon disulfide (CS2) on women employed in viscose industry. Med Pr. 2018;69(3):329–35

10. Bedi R. Evaluation of occupational environment in two textile plants in Northern India with specific reference to noise. Ind Health. 2006 Jan;44(1):112–6

11. Sellappa S, Prathyumnan S, Joseph S, Keyan KS, Balachandar V. Genotoxic effects of textile printing dye exposed workers in India detected by micronucleus assay. Asian Pac J Cancer Prev. 2010;11(4):919–22

12. Singh MB, Fotedar R, Lakshminarayana J. Occupational morbidities and their association with nutrition and environmental factors among textile workers of desert areas of Rajasthan, India. J Occup Health. 2005 Sep;47(5):371–7

13. Pinney C. On living in the kal(i)yug: Notes from Nagda, Madhya Pradesh. Contributions to Indian Sociology. 1999 Feb 1;33(1–2):77–106

14. Central Pollution Control Board. Comprehensive Environmental Assessment of Industrial Clusters [Internet]. New Delhi, India: Ministry of Environment and Forests; 2009 Dec [cited 2021 Aug 9]. (Ecological Impact Assessment Series). Report No.: EIAS/5/2009–2010. Available from: https://cpcb.nic.in/displaypdf.php?id=Q1BBL05ld0l0ZW1fMTUyX0ZpbmFsLUJvb2tfMi5wZGY=

15. Order of the National Green Tribunal regarding pollution by Grasim Chemical Division, Birlagram, Ujjain district, Madhya Pradesh [Internet]. India Environment Portal; 2020 [cited 2021 Feb 9]. Available from: http://www.indiaenvironmentportal.org.in/files/file/Nagada-Grasim-industry-NGT-order-April7-2021.pdf

16. Madhya Pradesh Population 2011-2020 [Internet]. [cited 2020 Aug 3]. Available from: https://www.census2011.co.in/census/state/madhya+pradesh.html

17. States Uts - Madhya Pradesh - Know India: National Portal of India [Internet]. [cited 2020 Aug 3]. Available from: https://knowindia.gov.in/states-uts/madhya-pradesh.php

18. Madhya Pradesh Presentation and Economy Growth Report | IBEF [Internet]. [cited 2021 Aug 13]. Available from: https://www.ibef.org/states/madhya-pradesh-presentation

19. Br.MSME-Development Institute. Brief Industrial Profile of Ujjain District Madhya Pradesh [Internet]. Rewa, Madhya Pradesh: Ministry of MSME, Govt. of India; [cited 2021 Aug 13] p. 14. Available from: http://dcmsme.gov.in/old/dips/Ujaain.pdf

20. Report on Prevention & Control of pollution in River Chambal An Action Plan for Rejuvenation [Internet]. Ujjain (M.P.): Regional Office M.P. Pollution control Board; 2019 [cited 2021 Aug 13]. Available from: http://www.mppcb.mp.gov.in/proc/poll-st-A-%20PLAN%2019/NGT%20Chambal.pdf

21. Reddy PB, Baghel BS. Impact of Industrial Wastewaters on the Physicochemical Characteristics of Chambal River at Nagda, M. P., India. Nature Environment and Pollution Technology. 2010;9(3):8

22. Kuo H-W, Lai J-S, Lin M, Su E-S. Effects of exposure to carbon disulfide (CS2) on electrocardiographic features of ischemic heart disease among viscose rayon factory workers. International Archives of Occupational and Environmental Health. 1997 Nov 1;70(1):61–6

23. Krishnan TN, Poulose S. Response rate in industrial surveys conducted in India: Trends and implications. IIMB Management Review. 2016 Jun 1;28(2):88–97

24. Martin-Olmedo P, Hams R, Santoro M, Ranzi A, Hoek G, de Hoogh K, et al. Environmental and health data needed to develop national surveillance systems in industrially contaminated sites. Epidemiol Prev. 2018 Dec;42(5-6S1):11–20

25. Sharan M, Yadav AK, Singh MP, Agarwal P, Nigam S. A mathematical model for the dispersion of air pollutants in low wind conditions. Atmospheric Environment. 1996 Apr 1;30(8):1209–20

26. National ambient air quality standards [Internet]. New Delhi: Central Pollution Control Board; 2009 [cited 2021 Sep 30]. Available from: https://cpcb.nic.in/openpdffile.php?id=UHVibGljYXRpb25GaWxlLzYzMF8xNDU3NTA2Mjk1X1B1YmxpY2F0aW9uXzUxNF9haXJxdWFsaXR5c3RhdHVzMjAwOS5wZGY=

27. SESDPROC-301-R3, Groundwater Sampling procedure, U.S. EPA, 2013 [Internet]. Georgia: U.S. EPA; 2013 [cited 2021 Sep 24]. Available from: https://www.epa.gov/sites/default/files/2015-06/documents/Groundwater-Sampling.pdf

28. Martin TD, Brockhoff CA, Creed JT, EMMC Methods Work Group. Method 200.7, Revision 4.4: Determination of Metals and Trace Elements in Water and Wastes by Inductively Coupled Plasma-Atomic Emission Spectrometry [Internet]. Cincinnati: US Environmental Protection Agency; 1994 [cited 2021 Sep 30]. Available from: https://www.epa.gov/sites/default/files/2015-08/documents/method_200-7_rev_4-4_1994.pdf

29. Djurić D, Surducki N, Berkes I. Iodine-azide test on urine of persons exposed to carbon disulphide. Br J Ind Med. 1965 Oct;22(4):321–3

30. Vasak V, Vanecek M, Kimmelova B. [ASSESSMENT OF THE EXPOSURE OF WORKERS IN AREAS CONTAMINATED BY CARBON DISULFIDE VAPORS. II. USE OF THE IODINE AZIDE REACTION FOR THE DETECTION AND ESTIMATION OF SULFIDE METABOLITES IN THE URINE]. Prac Lek. 1963 May;15:145–9

31. Rosier J, Billemont G, Van Peteghem C, Vanhoorne M, Grosjean R, Van de Walle A. Relation between the iodine azide test and the TTCA test for exposure to carbon disulphide. Br J Ind Med. 1984 Aug;41(3):412–6

32. National Centre for Disease Control. National Programme for Prevention and Control of Diabetes, Cardiovascular Disease and Stroke: Training Module for Medical Officers for Prevention, Control and Population Level Screening of Hypertension, Diabetes and Common Cancer (Oral, Breast & Cervical) [Internet]. New Delhi: Ministry of Health and Family Welfare, Government of India.; 2017 [cited 2021 Aug 6]. Available from: http://nhsrcindia.org/sites/default/files/Module%20for%20MOs%20for%20Prevention%2CControl%20%26%20PBS%20of%20Hypertension%2CDiabetes%20%26%20Common%20Cancer.pdf

33. Unger T, Borghi C, Charchar F, Khan NA, Poulter NR, Prabhakaran D, et al. 2020 International Society of Hypertension Global Hypertension Practice Guidelines. Hypertension. 2020 Jun 1;75(6):1334–57

34. Kruse A, Borch-Johnsen K, Pedersen LM. Cerebral Damage Following a Single High Exposure to Carbon Disulphide. Occupational Medicine. 1982 Jan 1;32(1):44–5

35. Ku M-C, Huang C-C, Kuo H-C, Yen T-C, Chen C-J, Shih T-S, et al. Diffuse White Matter Lesions in Carbon Disulfide Intoxication: Microangiopathy or Demyelination. European Neurology. 2003;50(4):220–4

36. De Fruyt F, Thiery E, De Bacquer D, Vanhoorne M. Neuropsychological Effects of Occupational Exposures to Carbon Disulfide and Hydrogen Sulfide. null. 1998 Jul 1;4(3):139–46

37. Krstev S, Peruničić B, Farkić B, Banićević R. Neuropsychiatric Effects in Workers with Occupational Exposure to Carbon Disulfide. Journal of Occupational Health. 2003 Mar 1;45(2):81– 7

38. Cassitto MG, Camerino D, Imbriani M, Contardi T, Masera L, Gilioli R. Carbon disulfide and the central nervous system: a 15-year neurobehavioral surveillance of an exposed population. Environ Res. 1993 Nov;63(2):252–63

39. International Programme on Chemical Safety, editor. Carbon disulfide [Internet]. Geneva: World Health Organization; 2002 [cited 2021 Nov 24]. 42 p. (Concise international chemical assessment document). Available from: https://www.who.int/ipcs/publications/cicad/en/cicad46.pdf

40. Putz-Anderson V, Albright BE, Lee ST, Johnson BL, Chrislip DW, Taylor BJ, et al. A behavioral examination of workers exposed to carbon disulfide. Neurotoxicology. 1983;4(1):67–77

41. Ganguli M, Ratcliff G, Chandra V, Sharma S, Gilby J, Pandav R, et al. A hindi version of the MMSE: The development of a cognitive screening instrument for a largely illiterate rural elderly population in india. International Journal of Geriatric Psychiatry. 1995 May 1;10(5):367–77

42. Tiwari RR, Raghvan S, Tripathi S. Cardiological and neurological health effects in viscose rayon workers exposed to carbon disulphide. 2012. (Unpublished manuscript)

43. Pradhan B, Nagendra H. Normative data for the digit-letter substitution task in school children. Int J Yoga. 2009 Jul;2(2):69–72

44. Natu M, Agarwal A. Testing of stimulant effects of coffee on the psychomotor performance: an exercise in clinical pharmacology. Indian Journal of Pharmacology. 1997;29:11

45. Fink HA, Hemmy LS, MacDonald R, Carlyle MH, Olson CM, Dysken MW, et al. Cognitive Outcomes After Cardiovascular Procedures in Older Adults: A Systematic Review [Internet]. Rockville (MD): Agency for Healthcare Research and Quality (US); 2014 [cited 2021 Nov 24]. (AHRQ Technology Assessments). Available from: http://www.ncbi.nlm.nih.gov/books/NBK285350/

46. Kennedy DO, Scholey AB. Glucose administration, heart rate and cognitive performance: effects of increasing mental effort. Psychopharmacology. 2000 Mar 1;149(1):63–71

47. Williams MA, LaMarche JA, Alexander RW, Stanford LD, Fielstein EM, Boll TJ. Serial 7s and Alphabet Backwards as brief measures of information processing speed. Archives of Clinical Neuropsychology. 1996 Jan 1;11(8):651–9

48. Corrigan JD, Hinkeldey NS. Relationships between Parts A and B of the Trail Making Test. Journal of Clinical Psychology. 1987 Jul 1;43(4):402–9

49. Ashendorf L, Horwitz JE, Gavett BE. Abbreviating the Finger Tapping Test. Archives of Clinical Neuropsychology. 2015 Mar 1;30(2):99–104

50. Aggarwal AN, Agarwal R, Dhooria S, Prasad KT, Sehgal IS, Muthu V, et al. Joint Indian Chest Society-National College of Chest Physicians (India) guidelines for spirometry. Lung India [Internet]. 2019 Apr;36(Supplement):S1–35. Available from: https://www.ncbi.nlm.nih.gov/pubmed/31006703

51. Sandau KE, Funk M, Auerbach A, Barsness GW, Blum K, Cvach M, et al. Update to Practice Standards for Electrocardiographic Monitoring in Hospital Settings: A Scientific Statement From the American Heart Association. Circulation. 2017 Nov 7;136(19):e273–344

52. Hampton J, Hampton J. The ECG Made Easy. 9th ed. Elsevier; 2019. (Made Easy)

53. Best practice in phlebotomy and blood collection [Internet]. WHO Best Practices for Injections and Related Procedures Toolkit. Geneva: World Health Organization; 2010 [cited 2021 Sep 24]. Available from: https://www.ncbi.nlm.nih.gov/books/NBK138496/

54. ICMR-National Institute of Medical Statistics. National Guidelines for Data Quality in Surveys [Internet]. New Delhi: ICMR-National Institute of Medical Statistics; 2021 [cited 2021 Sep 27]. Available from: https://ndqf.in/wp-content/uploads/2021/07/National-Guidelines-for-DATA-QUALITY-in-Surveys.pdf

55. Baker I. Rayon. In: Baker I, editor. Fifty Materials That Make the World [Internet]. Cham: Springer International Publishing; 2018. p. 195–7. Available from: https://doi.org/10.1007/978-3-319-78766-4_37

56. Jiang X, Bai Y, Chen X, Liu W. A review on raw materials, commercial production and properties of lyocell fiber. Journal of Bioresources and Bioproducts. 2020 Feb 1;5(1):16–25

57. Gelbke H-P, Göen T, Mäurer M, Sulsky SI. A review of health effects of carbon disulfide in viscose industry and a proposal for an occupational exposure limit. Crit Rev Toxicol. 2009 Oct;39 Suppl 2:1–126

58. Sieja K, Mach-Szczypiński J von, Mach-Szczypiński J von. Health effect of chronic exposure to carbon disulfide (CS 2) on women employed in viscose industry. Med Pr. 2018 May 22;69(3):329– 35

59. Chang S-J, Chen C-J, Shih T-S, Chou T-C, Sung F-C. Risk for hypertension in workers exposed to carbon disulfide in the viscose rayon industry. Am J Ind Med. 2007 Jan;50(1):22–7

60. Chang S-J, Shih T-S, Chou T-C, Chen C-J, Chang H-Y, Chen P-C, et al. Electrocardiographic abnormality for workers exposed to carbon disulfide at a viscose rayon plant. J Occup Environ Med. 2006 Apr;48(4):394–9

61. Takebayashi T, Nishiwaki Y, Uemura T, Nakashima H, Nomiyama T, Sakurai H, et al. A Six Year Follow up Study of the Subclinical Effects of Carbon Disulphide Exposure on the Cardiovascular System. Occupational and Environmental Medicine. 2004;61(2):127–34

62. Corsi G, Maestrelli P, Picotti G, Manzoni S, Negrin P. Chronic peripheral neuropathy in workers with previous exposure to carbon disulphide. Br J Ind Med. 1983 May;40(2):209–11

63. Ruijten MW, Sallé HJ, Verberk MM, Muijser H. Special nerve functions and colour discrimination in workers with long term low level exposure to carbon disulphide. Br J Ind Med. 1990 Sep;47(9):589–95

64. Godderis L, Braeckman L, Vanhoorne M, Viaene M. Neurobehavioral and clinical effects in workers exposed to CS (2). International journal of hygiene and environmental health. 2006 Apr 1;209:139–50

65. Spyker DA, Gallanosa AG, Suratt PM. Health effects of acute carbon disulfide exposure. J Toxicol Clin Toxicol. 1982 Mar;19(1):87–93

66. White CW, Martin JG. Chlorine gas inhalation: human clinical evidence of toxicity and experience in animal models. Proc Am Thorac Soc. 2010 Jul;7(4):257–63

67. Lewis RJ, Copley GB. Chronic low-level hydrogen sulfide exposure and potential effects on human health: a review of the epidemiological evidence. Crit Rev Toxicol. 2015 Feb;45(2):93–123

68. Wu Y, Li R, Cui L, Meng Y, Cheng H, Fu H. The high-resolution estimation of sulfur dioxide (SO2) concentration, health effect and monetary costs in Beijing. Chemosphere. 2020 Feb 1;241:125031

69. Konstantinova E, Burachevskaya M, Mandzhieva S, Bauer T, Minkina T, Chaplygin V, et al. Geochemical transformation of soil cover and vegetation in a drained floodplain lake affected by long-term discharge of effluents from rayon industry plants, lower Don River Basin, Southern Russia. Environmental Geochemistry and Health [Internet]. 2020 Aug 6; Available from: https://doi.org/10.1007/s10653-020-00683-3

70. Kumar S, Toppo S, Kumar A, Tewari G, Beck A, Bachan V, et al. Assessment of heavy metal pollution in groundwater of an industrial area: a case study from Ramgarh, Jharkhand, India. null. 2020 Oct 6;1–23

71. Shaji E, Gómez-Alday JJ, Hussein S, Deepu TR, Anilkumar Y. Salinization and Deterioration of Groundwater Quality by Nitrate and Fluoride in the Chittur Block, Palakkad, Kerala. Journal of the Geological Society of India. 2018 Sep 1;92(3):337–45

72. Ding C-Q, Li K-R, Duan Y-X, Jia S-R, Lv H-X, Bai H, et al. Study on community structure of microbial consortium for the degradation of viscose fiber wastewater. Bioresources and Bioprocessing. 2017 Jul 10;4(1):31

73. Vanhoorne M, Berge LVD, Devreese A, Tijtgat E, Poucke LV, Peteghem CV. SURVEY OF CHEMICAL EXPOSURES IN A VISCOSE RAYON PLANT. The Annals of Occupational Hygiene. 1991 Dec 1;35(6):619–31

74. Briffa J, Sinagra E, Blundell R. Heavy metal pollution in the environment and their toxicological effects on humans. Heliyon. 2020 Sep 8;6(9):e04691–e04691

75. Employment Generated Through Textile Sector [Internet]. [cited 2021 Dec 23]. Available from: https://pib.gov.in/pib.gov.in/Pressreleaseshare.aspx?PRID=1575203

76. The Changing Markets Foundation. Pollution and disease: report reveals disastrous impact of the viscose giant behind high street brands [Internet]. 2018 [cited 2021 Aug 13]. Available from: http://changingmarkets.org/wp-content/uploads/2018/02/Dirty-Fashion-2018-press-release-FINAL.pdf

77. Grasim Industries Limited. Nagda Environment Clearance Compliance Report (Oct 20-Mar 21) [Internet]. 2021 [cited 2021 Nov 24]. Available from: https://www.grasim.com/Upload/PDF/nagda-environment-clearance-compliance-report-oct-20-mar-21.pdf

